# Gene-environment interactions for Parkinson’s disease

**DOI:** 10.1101/2023.06.15.23291423

**Authors:** Alexandra Reynoso, Roberta Torricelli, Benjamin Meir Jacobs, Jingchunzi Shi, Stella Aslibekyan, Lucy Kaufmann, Alastair J Noyce, Karl Heilbron

## Abstract

**Importance:** Parkinson’s disease (PD) is a neurodegenerative disorder with complex aetiology. Multiple genetic and environmental factors have been associated with PD, but most PD risk remains unexplained.

**Objective:** The aim of this study was to test for statistical interactions between PD-related genetic and environmental exposures/phenotypic traits in the 23andMe, Inc. research dataset.

**Design:** Nested cross-sectional case-control study.

**Setting:** Population-based cohort.

**Participants:** PD subjects were recruited to join the 23andMe, Inc study population in collaboration with the Michael J. Fox Foundation and other PD patient advocacy groups, and/or via online surveys. Participants that reported a change or uncertainty in diagnosis during follow-up were excluded. Controls were recruited from 23andMe participants that did not report a diagnosis of PD at entry or on subsequent follow-up surveys.

**Exposures:** Using a validated PD polygenic risk score (PRS) and common PD-associated variants in the *GBA* gene, we explored interactions between genetic susceptibility factors and phenotypic traits: body mass index (BMI), type 2 diabetes (T2D), tobacco use, caffeine consumption, pesticide exposure, head injury, and physical activity (PA).

**Main Outcomes and Measures:** Self-reported PD case/control status.

**Results:** The dataset contained 18,819 PD cases (40.2% female) and 545,751 controls (55.7% female). The average age of PD cases and controls was 73.1 and 73.0 years, respectively (SD_PD_ = 10.8 years, SD_control_ = 10.8 years). In models without gene-by-environment interactions, we observed that higher BMI, T2D, caffeine consumption, and tobacco use were associated with lower odds of PD, while head injury, pesticide exposure, and *GBA* carrier status were associated with increased odds. We observed no significant association between PA and PD. PRS was associated with increased odds of PD and there was statistical evidence for an interaction between PRS and BMI, PRS and T2D, PRS and PA, and PRS and tobacco use (p=4.314E-4; p=6.502E-8; p=8.745E-5, p=2.236E-3, respectively). Whilst BMI and tobacco use were associated with lower odds of PD regardless of the extent of individual genetic liability, the direction of the relationship between odds of PD and T2D as well as PA, varied depending on PRS.

**Conclusions and Relevance:** We provide preliminary evidence that associations between phenotypic traits and PD may be modified by genotype.

## INTRODUCTION

Parkinson’s disease (PD) may be the fastest growing neurological disorder worldwide, with a prevalence of 1-2% in the over 60 population (1). It is characterised by chronic, progressive neuronal loss and intracellular alpha-synuclein inclusions (Lewy bodies). The genetic architecture of PD involves contributions from both common, modest effect-size alleles and rarer, monogenic forms identified via linkage analyses of affected families such as *SNCA, PINK1, PARK7*, and *PRKN* (2). Over the last decade, genome-wide association studies (GWAS) have identified susceptibility loci involved in complex disease, broadening our understanding of the genetic basis of PD (3,4). Among these studies, variants of *GBA* and *LRRK2* genes were recognised as the most common risk factors for PD (2,5).

Calculation of polygenic risk scores (PRS) have allowed for more in depth assessments of the cumulative effects of independent risk loci at an individual level, explaining 16-36% of PD heritability (4). This information on genetic risk can be further characterised in conjunction with modifiable phenotypic risk factors identified through epidemiological studies, thereby advancing our understanding of complex disease aetiology (6).

Previous studies have examined interactions between *LRRK2* and various environmental exposures such as tobacco, black tea consumption and NSAID use, all of which were shown to be associated with a decreased risk of disease penetrance (7,8). Other studies have examined evidence for interactions between PD PRS and common health risk factors, such as diabetes, alcohol and tobacco consumption, which revealed a complex genetic aetiology with variable evidence for environmental interactions (9). In the current project, our goal was to test whether interactions between genetic (PD PRS and *GBA* variants) and putative environmental and/or phenotypic determinants influence PD risk.

## SUBJECT AND METHODS

### Participants

The dataset consisted of customers of 23andMe, Inc., a direct-to-consumer company. Informed written consent to participate in research was obtained from all participants. 23andMe’s human subjects protocol was reviewed and approved by Ethical & Independent Review Services, an AAHRPP-accredited institutional review board (IRB).

### IRB Statement

Participants provided informed consent and volunteered to participate in the research online, under a protocol approved by the external AAHRPP-accredited IRB, Ethical & Independent (E&I) Review Services. As of 2022, E&I Review Services is part of Salus IRB (https://www.versiticlinicaltrials.org/salusirb).

### Parkinson’s disease status

PD subjects were recruited to join the study population in collaboration with the Michael J. Fox Foundation and other PD patient advocacy groups from 2009 onwards. In addition, individuals with PD were identified via online surveys. Research participants that self-reported a diagnosis of PD were targeted to receive additional follow-up surveys regarding their symptoms, risk factors, lifestyle habits and lifetime environmental exposures. We excluded cases that reported a change in diagnosis or uncertainty about their diagnosis on follow-up.

Controls were recruited from the pool of participants that did not report a diagnosis of PD at entry or on subsequent follow-up surveys.

### Phenotypic factors

We prioritised seven phenotypes that have been repeatedly reported in previous PD epidemiological studies as modifiable environmental exposures or comorbidities (10,11): type 2 diabetes (T2D), tobacco use, caffeine consumption, body mass index (BMI), pesticide exposure, head injury, and physical activity (PA). These phenotypes were extracted from self-reported survey answers. T2D was recorded as the presence of a previous diagnosis (yes/no). BMI was calculated as mass (kg) divided by height squared (m^2^) and quantile normalised separately in males and females. Tobacco consumption was determined by a smoking history of at least 100 cigarettes (yes/no). Caffeine consumption was measured as daily milligrams of caffeine from any of the following: coffee, tea, soda, or energy drinks. Caffeine consumption was then transformed by log_10_(x+75) to create a more normally distributed variable. Pesticide exposure was defined as any monthly frequency of home, garden, or pest-related pesticide use (yes/no). Head injury history was recorded as the result of sporting activities, falls, violence, car accidents, or other accidents (yes/no). Physical activity was measured by determining the amount of times per week a participant engaged in physical activity for longer than 30 minutes. All phenotypes were recorded cross-sectionally along with PD case-control status. As such, the analysis is limited to exploring interactions in cross-sectional data rather than examining temporal associations.

### Genotyping

DNA extraction and genotyping were performed on saliva samples by Clinical Laboratory Improvement Amendments-certified and College of American Pathologists-accredited clinical laboratories of Laboratory Corporation of America.□ Samples were genotyped on one of five genotyping platforms. The V1 and V2 platforms were variants of the Illumina HumanHap550+ BeadChip and contained a total of approximately 560,000 SNPs, including about 25,000 custom SNPs selected by 23andMe. The V3 platform was based on the Illumina OmniExpress + BeadChip and contained a total of about 950,000 SNPs and custom content to improve the overlap with our V2 array.□The V4 platform is a fully custom array and includes a lower redundancy subset of V2 and V3 SNPs with additional coverage of lower-frequency coding variation, and about 570,000 SNPs.□ The V5 platform is an Illumina Infinium Global Screening Array of about 640,000 SNPs supplemented with about 50,000 SNPs of custom content.□Samples had minimum call rates of 98.5%.

### Imputation

We imputed variants using two separate imputation reference panels. First, we used the Human Reference Consortium (HRC) imputation reference panel (12) (32,488 samples, 39,235,157 SNPs). Second, we combined the May 2015 release of the 1000 Genomes Phase 3 haplotypes (13) with the UK10K imputation reference panel (14) to create a single unified panel. To do this, multiallelic sites with N alternate alleles were split into N separate biallelic sites. We then removed any site whose minor allele appeared in only one sample. For each chromosome, we used Minimac3 (15) to impute the reference panels against each other, reporting the best-guess genotype at each site. This gave us calls for all samples over a single unified set of variants. We then joined these together to get, for each chromosome, a single file with phased calls at every site for 6,285 samples. Throughout, we treated structural variants and small indels in the same way as SNPs. In preparation for imputation, we split each chromosome of the reference panel into chunks of no more than 300,000 variants, with overlaps of 10,000 variants on each side. We used a single batch of 10,000 individuals to estimate Minimac3 imputation model parameters for each chunk.

To generate phased participant data for the V1 to V4 platforms, we used an internally developed tool, Finch, which implements the Beagle graph-based haplotype phasing algorithm (16), modified to separate the haplotype graph construction and phasing steps. Finch extends the Beagle model to accommodate genotyping error and recombination, in order to handle cases where there are no consistent paths through the haplotype graph for the individual being phased. We constructed haplotype graphs for all participants from a representative sample of genotyped individuals, and then performed out-of-sample phasing of all genotyped individuals against the appropriate graph.

We then built a merged imputation dataset by combining the two sets of imputed data. We applied a simple merging rule: if a variant was imputed in the HRC panel, the HRC imputed results were included in the merged imputed dataset. For the remaining variants not present in HRC (all indels and structural variants, for example), the 1KG/UK10K imputed results were added to the merged dataset. In total, the merged imputed dataset contained 64,439,130 variants.

### *GBA* variants

We aggregated three known *GBA* PD risk-associated variants E326K (rs2230288), T369M (rs75548401) and N370S (rs76763715) into a single binary variable (referred to hereafter as “*GBA* carrier status”) denoting whether an individual was a carrier for any variant. Following the methods of previous groups (17); variants E326K, T369M and N370S were combined into a single *GBA* variable due to their similar effects on GCase activity in humans, reducing it by 18–46% on average (18).

### PRS calculation

To select SNPs for inclusion into the PRS, we used summary statistics from the most recently published PD GWAS excluding 23andMe participants (4). We restricted the analysis to common (minor allele frequency [MAF] >1%), biallelic, autosomal, non-palindromic SNPs. SNPs were selected using the ‘clumping-and-thresholding (C+T)’ approach. We used all possible combinations of 5 clumping thresholds and 11 P value thresholds to generate 55 PRSs. SNPs were LD-clumped using PLINK (version 1.9) with a clumping distance of 250kb and five clumping R^2^ thresholds: 0.1, 0.2, 0.4, 0.6, and 0.8. The 503 European-ancestry samples from the 1,000 Genomes project were used as the LD reference. Using P values for each SNP’s association with PD, we filtered SNPs using 11 P value thresholds: 5e-8, 5e-5, 5e-4, 0.005, 0.05, 0.1, 0.2, 0.4, 0.6, 0.8, and 1. Effect size estimates for the association of each variant with PD were obtained from the GWAS beta coefficient, representing the per-allele log odds ratio for PD.

We constructed 55 PRSs for 23andMe research participants as the weighted sum of risk allele counts for the SNPs selected in each of the 55 C+T profiles. We matched SNPs from the PD GWAS to 23andMe SNPs using the CPRA (chromosome, position, reference allele, alternative allele) format and excluding unmatched variants (4). We harmonised SNP effect size estimates following the variant harmonisation schema in (19). To ensure good SNP quality, we removed SNPs with imputation R^2^ < 0.5 or a difference in MAF > 30% between the PD GWAS variant and the 23andMe variant. For each of the 55 C+T profiles, PRS values were calculated for each individual, *i*, as 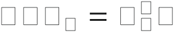, where *β* is an □ X *1* vector of harmonised weights for the □ selected SNPs and □_□_ is the corresponding □ X *1* vector of imputed dosages of the □ selected SNPs in individual □. Lastly, we standardised the PRS to have mean 0 and standard deviation 1.

In order to identify the best-performing PRS, we split our dataset into a 30% test set and a 70% held-out set. In the test set we performed 55 logistic regressions using PD as the dependent variable, 1 of the 55 PRSs as the independent variable, and the following covariates: age, sex, household income (inferred from zip code), and five principal components of genetic ancestry. We selected the PRS with the largest McFadden R^2^ value for use in the 70% held-out set. The best-performing PRS used a clumping R^2^ of 0.8 and a p-value threshold of 0.4. The corresponding C+T profile contained 603,976 SNPs prior to harmonisation and 593,886 SNPs (98.3%) after harmonisation.

### Age-matched cross-sectional case-control datasets

For each of the seven phenotypic factors, we constructed age-matched, PD case-control datasets; including all individuals from the 70% held-out set with available data for the phenotypic factor. Specifically, we divided PD cases into 20 evenly populated age bins; determined the number of controls that fell into each corresponding age bin; divided the number of controls in a bin by the number of corresponding cases; determined the minimum number of available controls per case across all bins; and randomly selected this minimum number of age-matched controls for each case. We also created an age-matched “full dataset” containing all PD cases regardless of whether data was available for the phenotypic factors. We used the full dataset for all analyses that did not use data from any of the seven phenotypic factors. These case-control datasets only contained individuals with predominantly European ancestry since the PRS was derived from a European ancestry GWAS (20). Supplementary Table 1 provides information on the sample size for each case-control dataset.

### Statistical analysis

All interaction tests were performed using logistic regression with PD status as the dependent variable and the following covariates: age, sex, and five principal components of genetic ancestry. For each phenotypic factor, we modelled the effect of the phenotypic factor, the PRS, and the interaction between the phenotypic factor and the PRS in the relevant age-matched dataset. We repeated these analyses but substituted *GBA* carrier status for the PRS. To isolate phenotypic associations, we also ran models without interactions or genetic variables. We used a Bonferroni adjusted p-value of 3.6E-3 (0.05/14 tests, seven phenotypic factors multiplied by two genetic factors) as the threshold for statistical significance for the interaction tests.

## RESULTS

To test for gene-by-environment interactions affecting PD risk, we constructed age-matched, case-control datasets derived from the 23andMe research database (Methods, Table 1). The full dataset contained 18,819 PD cases (40.2% female) and 545,751 controls (55.7% female). PD cases and controls had an average age of 73.1 and 73.0 years, respectively (SD_PD_ = 10.8 years, SD_control_ = 10.8 years). We tested two genetic factors: a PRS derived from the largest published PD GWAS (4) and *GBA* carrier status (PD = 7.6% carriers, control = 4.5% carriers). We tested seven phenotypic factors: physical activity, BMI, T2D, tobacco use, pesticide exposure, head injury, and caffeine intake (Table 2). Because data availability varied across phenotypic factors, we constructed separate age-matched datasets for each phenotypic factor (median N_PD_ = 14,692, median N_control_ = 520,056). Age and sex distributions were similar across all data sets (PD mean age: 71.5 -72.9 years, PD percentage female: 40.3% - 43.8%).

**Table 1:**

|  | Cases |  | Controls |  | P |
| --- | --- | --- | --- | --- | --- |
|  |  | N |  | N |  |
| Age, mean (SD), years | 73.1 (10.8) | 18,819 | 73.0 (10.8) | 545,751 | $1.9 \times 10^{-1}$ |
| Female, no. (%) | 7,599 (40.2%) | 18,819 | 303,979 (55.7%) | 545,751 | $< 1 \times 10^{-300}$ |
| PRS, mean (SD) | 0.3 (1.0) | 18,819 | -0.01 (1.0) | 545,751 | $< 1 \times 10^{-300}$ |
| *GBA* carrier status, no. (%) | 1,438 (7.6%) | 18,819 | 23,793 (4.5%) | 545,751 | $1.3 \times 10^{-80}$ |
| Physical activity, mean (SD) | 3.1 (2.6) | 14,695 | 3.1 (2.6) | 602,495 | $2.6 \times 10^{-1}$ |
| Q-norm body mass index, mean (SD), kg/m<sup>2</sup> | -0.2 (1.1) | 16,843 | 0.01 (1.0) | 555,819 | $4.6 \times 10^{-193}$ |
| Type 2 diabetes, no. (%) | 1,699 (11.5%) | 16,425 | 64,090 (11.1%) | 574,875 | $9.5 \times 10^{-9}$ |
| Tobacco use, no. (%) | 6,547 (39.0%) | 16,776 | 251,252 (46.8%) | 536,832 | $1.9 \times 10^{-106}$ |
| Pesticide exposure, no. (%) | 2,953 (42.1%) | 7,022 | 88,186 (38.1%) | 231,726 | $5.5 \times 10^{-11}$ |
| Head injury, no. (%) | 3,137 (41.0%) | 7,652 | 28,117 (33.4%) | 84,172 | $2.4 \times 10^{-22}$ |
| Caffeine | 2.3 (0.3) | 10,364 | 2.4 (0.4) | 93,276 | $6.2 \times 10^{-168}$ |
| consumption,<br>mean (SD), mgs |  |  |  |  |  |

**Table 2:**

|  | OR | 95% CI | p-value |
| --- | --- | --- | --- |
| Head Injury | 1.31 | 1.25 - 1.38 | 7.03x10<sup>-28</sup> |
| Pesticide Exposure | 1.16 | 1.11 - 1.22 | 8.17x10<sup>-10</sup> |
| Type 2 Diabetes | 0.86 | 0.82 - 0.91 | 1.97x10<sup>-8</sup> |
| Q-norm BMI | 0.79 | 0.78 - 0.80 | 2.48x10<sup>-190</sup> |
| Tobacco Use | 0.70 | 0.68 - 0.72 | 2.42x10<sup>-108</sup> |
| Caffeine Consumption | 0.43 | 0.41 - 0.46 | 1.80x10<sup>-167</sup> |
| Physical Activity | 0.99 | 0.99 - 1.00 | 0.111 |

Legend: OR - odds ratio, 95% CI - 95% confidence interval
Order: positive association, inverse association, null

In the regression models without interaction terms, we observed negative associations between PD and caffeine intake (OR = 0.43, 95% CI = 0.41 - 0.46, P = 1.797x10^−167^, N_PD_ = 10,364, N_controls_ = 93,276), tobacco use (OR = 0.70, 95% CI = 0.68 - 0.72, P = 2.417x10^−108^, N_PD_ = 16,776, N_controls_ = 536,832), BMI (OR = 0.79, 95% CI = 0.78 - 0.80, P = 2.484x10^−190^, N_PD_ = 16,843, N_controls_ = 555,819), and T2D (OR = 0.86, 95% CI = 0.82 - 0.91, P = 1.966x10^− 8^, N_PD_ = 16,425, N_controls_ = 574,875). We found positive associations between PD and pesticide exposure (OR = 1.16, 95% CI = 1.11 - 1.22, P = 8.168x10^−10^, N_PD_ = 7,022, N_controls_ = 231,726), head injury (OR = 1.31, 95% CI = 1.25 - 1.38, P = 7.027x10^−28^, N_PD_ = 7,652, N_controls_ = 84,172), PRS (OR per standard deviation = 1.41, 95% CI = 1.39 - 1.43, P < 1x10^− 300^, N_PD_ = 18,819, N_controls_ = 545,751), and *GBA* carrier status (OR = 1.73, 95% CI = 1.63 - 1.83, P = 2.072x10^−82^, N_PD_ = 18,864, N_controls_ = 528,192). There was no significant association between PD and physical activity (OR = 0.99, 95% CI = 0.99 - 1.00, P =1.113x10^−1^, N_PD_ = 14,695, N_controls_ = 602,495).

In the regression models with interaction terms, we found significant interactions between the PRS and T2D (OR = 0.87, 95% CI = 0.83 - 0.91, P = 6.502x10^−8^), BMI (OR = 0.97, 95% CI = 0.96 - 0.99, P = 4.314x10^−4^), PA (OR = 1.01, 95% CI = 1.01 - 1.02, P = 8.745x10^−5^) and tobacco use (OR = 0.95, 95% CI = 0.92 – 0.98, P = 2.236x10^−3^) (Table 3). No significant interactions were observed in the *GBA* analysis.

**Table 3:**
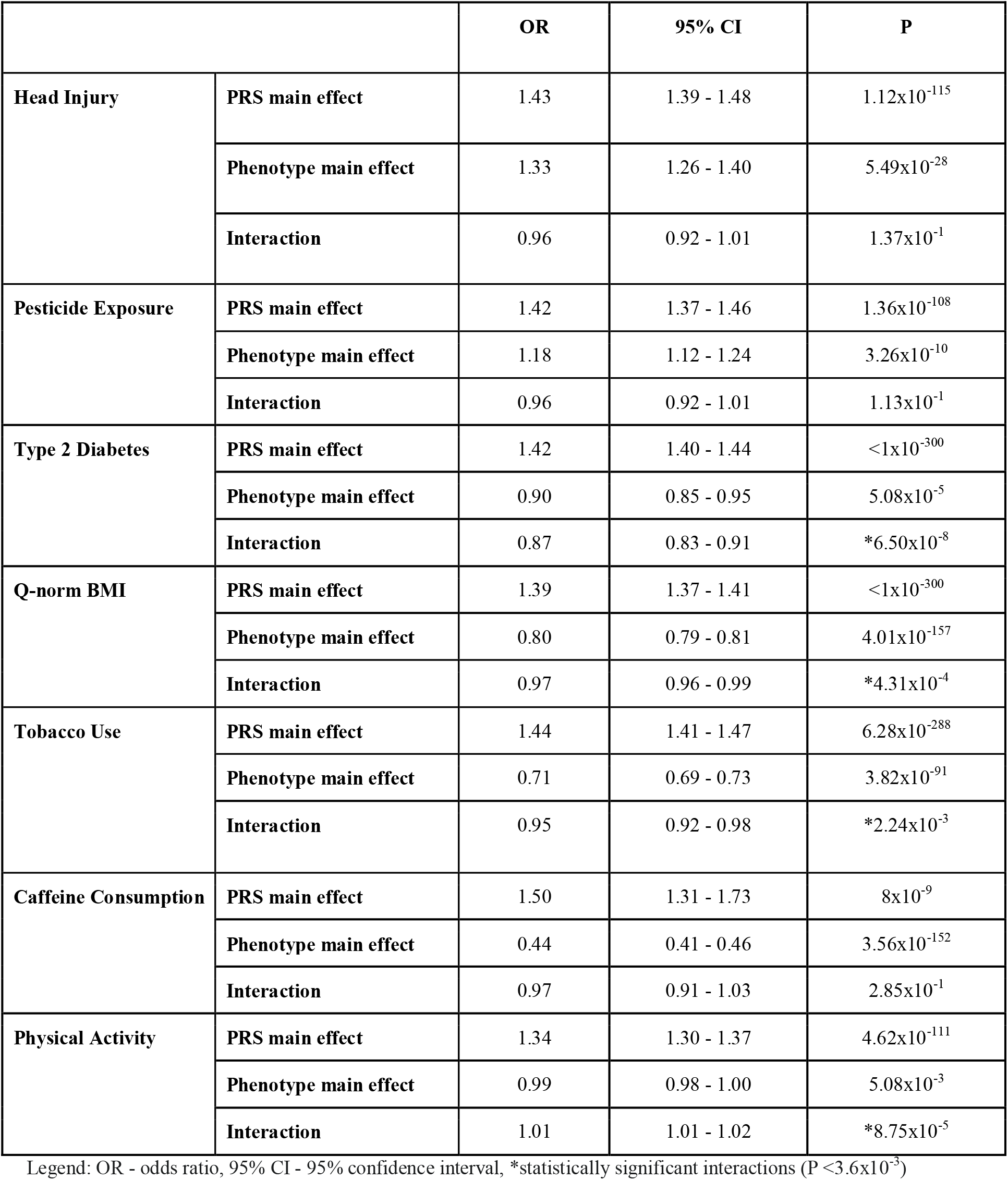

**Figure 1:**
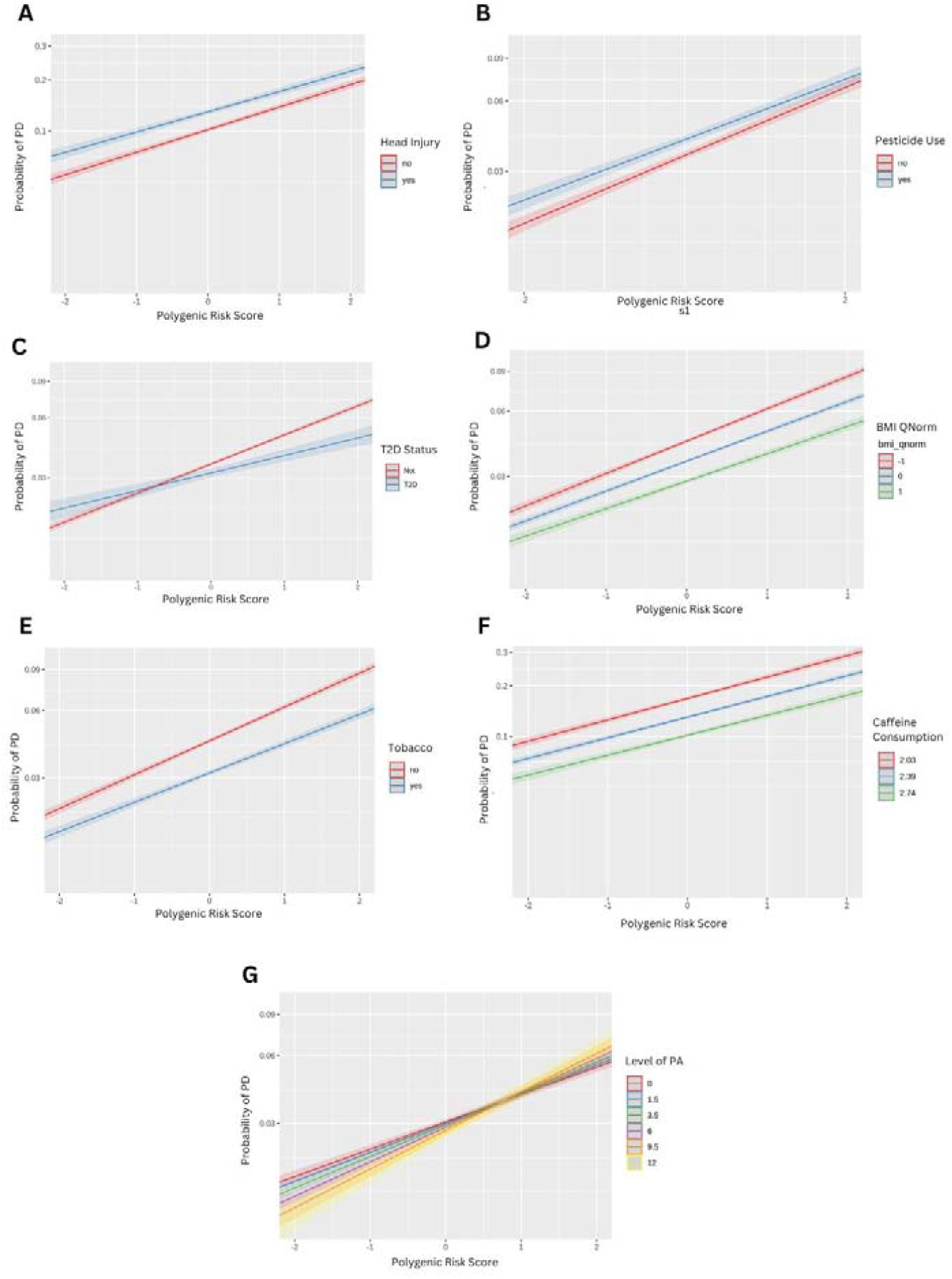
Marginal effects plots for the seven PRS-by-phenotype interaction models. Lines represent the fitted probability of PD for a given pair of PRS and phenotypic factor values, shaded areas represent 95% confidence intervals. The y-axis displays the fitted probability of PD on a logit-transformed scale to preserve a linear relationship with the PRS and phenotypic factors. The x-axis window spans ±2 standard deviations of PRS, covering ∼95% of individuals. The phenotypic factor values plotted are as follows: **(A)** presence or absence of head injury; **(B)** presence or absence of pesticide exposure; **(C)** presence or absence of a T2D diagnosis; **(D)** quantile-normalised body mass index at the sample mean (0), one standard deviation below the sample mean (-1), or one standard deviation above the sample mean (1); **(E)** presence or absence of tobacco use; **(F)** log_10_-transformed caffeine intake at the sample mean (2.39), one standard deviation below the sample mean (2.03), or one standard deviation above the sample mean (2.74); **(G)** number of 30-minute bouts of physical activity per week for all possible survey response options (0, 1.5, 3.5, 6, 9.5, or 12).

## DISCUSSION

In this large, cross-sectional case-control study, we observed evidence for statistical interactions between PRS and BMI, PRS and T2D, PRS and PA, and PRS and tobacco consumption on risk of PD. For individuals with a PRS value of 0 (the sample mean), a 1-SD increase in quantile-normalised BMI was associated with a 20% decrease in the odds of PD. This same change in BMI was associated with a 22% and 18% decrease in the odds of PD for individuals with PRS values 1 standard deviation above or below the mean, respectively. A similar interaction was observed for tobacco use. For individuals with a PRS value of 0, tobacco use was associated with a 29% decrease in the odds of PD. This varied from a 33% to a 26% decrease in the odds of PD for individuals with PRS values 1 standard deviation above or below the mean, respectively.

The interaction observed for T2D was more complicated. For individuals with a PRS value of 0, T2D was associated with a 10% decrease in the odds of PD. For individuals with a PRS value 1 SD below the mean, however, T2D was associated with a 3% increase in PD odds. This suggests a potential qualitative interaction between T2D and genotype on PD risk, which was similar to an interaction between PRS and T2D we previously reported in an analysis using UK Biobank data (21). Like T2D, PA was associated with increased or decreased PD risk depending on an individual’s PRS value. For individuals with a PRS value of 0, one 30-minute bout of PA per week was associated with a 1% decrease in the odds of PD. For individuals with a PRS value 1 SD above the mean, however, one 30-minute bout of PA per week was associated with a 0.5% increase in PD odds.

There remains a large proportion of PD risk that remains unexplained by genetic variation, environmental exposures, lifestyle factors, or comorbidities. Exploring interactions between genetic and non-genetic factors may ultimately yield insights into disease risk through follow-up investigation. However, interpreting such interactions is far from straightforward and so, rather than offering mechanistic insights, this study should be viewed as providing evidence for proof of principle (22). We observed four significant statistical interactions in the present analysis. For two phenotypic factors, BMI and tobacco, the inverse associations between phenotype and PD were attenuated in the presence of a higher genetic risk of PD and magnified in the presence of a lower genetic risk.

For T2D, the nature of the interaction was more complicated, reversing directions of association at lower levels of PRS. The relationship between T2D and PD has been extensively studied and there is increasing evidence for shared underlying pathways and mechanisms (23). Most prospective cohort studies that ascertained T2D prior to PD diagnosis endorse a modest increase in the risk of PD associated with T2D (24). However, cross-sectional and case-control studies often reveal inverse associations between T2D and PD, as seen in the present study. This raises the possibility of bias due to selective mortality, in study designs which are more susceptible to this type of bias (24). Evidence for T2D conveying an increased risk of PD in high-quality, prospective studies is supported by Mendelian randomisation (24). We previously showed an interaction between T2D and PRS using data from UK Biobank (21). In that prospective cohort study, the interaction suggested that T2D was a more potent risk factor in those with a lower genetic liability towards PD. As such, in both studies, the magnitude of the association was greatest in those with lower genetic liability.

Observational studies suggest that T2D may also contribute to more rapid disease progression (24, 25, 26). Drugs used to treat T2D are being widely repurposed and tested to see if they might modify the course of PD. Although the results presented here reflect PD risk rather than progression, they raise the possibility that genetic stratification could be important when recruiting patients to such trials in order to identify subgroups that will have the best response.

There exists compelling observational evidence for an inverse association between PA and PD (27, 28, 29, 30). Again, a serious challenge is unpicking reverse causality from a causal relationship. Individuals who have undertaken regular PA appear to be at reduced risk of PD, but it is also probable that in the early stages of disease PA reduces due to occult disease. We found no association between PA and PD in our regression model without an interaction effect. In our regression model with an interaction effect, however, a protective association was apparent with greater levels of PA in participants with lower genetic liability. Several PA-based intervention studies have already been conducted and have shown improvements in ‘off’ state UPDRS scores, hinting at possible disease modifying benefits (31). PA-focussed intervention studies are imminent for those at risk of future PD and the current results suggest that genetic stratification could be important in their design and interpretation.

Observational studies examining the association between BMI and PD are complex to interpret given the dynamic nature of BMI during the course of PD. Generally, a reduction in BMI is seen following a diagnosis of PD and may precede the diagnosis (32). This could be attributed to the disruption of normal homeostatic and hedonic mechanisms which result from neuroendocrine changes of disease pathogenesis (33). Given the possibility of reverse causation, a meta-analysis of prospective cohort studies that measured BMI prior to PD diagnosis demonstrated no overall association (34). Previous Mendelian randomisation studies further examined the association of genetically estimated BMI and liability towards PD, again concluding an inverse association that was not thought to be explained by survival bias (35, 36). The issue of survival bias is important when considering exposures that are associated with premature mortality and age-related outcomes such as PD, including higher BMI and T2D. Several of the intriguing inverse associations that have been reported for PD might be driven, in part or entirely, by survivor bias or other types of bias. The cross-sectional nature of the present study prevents us from generating conclusive insights about the direction and nature of the association between BMI and PD, but indicates that it may be modified by genotype.

An inverse association between smoking and PD risk has long been recognised. Epidemiological studies consistently highlight reduced odds of disease in individuals who smoke tobacco-containing cigarettes or are indirectly exposed to tobacco smoke (10, 37, 38, 39,40). In the present study, we not only observed the presence of an inverse association between tobacco consumption and odds of PD, but also a potential interaction between smoking and PRS such that the protective association with smoking was greatest in those at higher genetic risk.

Genetic stratification in future clinical trials seems inevitable and will help underpin a general shift towards precision medicine (41). Randomised controlled trials are an excellent way to examine causal relationships but are not always ethical and/or practical. Based on the current results, existing and planned clinical trials focused on repurposed drugs for T2D and non-drug interventions like PA, may benefit from genetic stratification of participants to build on the initial observations we report here (42,43).

A limitation of this study was that the definition of both PD cases and other related phenotypes relied on self-reporting. This type of data collection can lead to bias and inaccuracy but has been previously validated in terms of the phenotype reporting and the genetic data (44,45). Furthermore, the dataset lacks temporal information regarding the seven selected phenotypic traits and PD, and is cross-sectional rather than longitudinal in nature. In addition, the 23andMe study population is not a random sample of the overall population and results derived from this type of sampling may not be generalizable to individuals who are not well-represented. For example, our study only contained individuals of European descent. This means that the findings may not be generalizable and should be investigated in other ancestral groups (46). Variability associated with ancestral diversity could account for differences in genetic risk factors, susceptibility to the environmental exposures, as well as interactions between the two.

Our study is the first to systematically examine interactions between selected phenotypic traits and common genetic variation related to PD. Use of 23andMe data meant that sufficiently large sample sizes could be used to investigate interaction, but the findings and implications should be followed by further research, including the re-analysis of data from previously completed T2D drug trials in PD and PA trials in PD, stratified by genotype.

## Data Availability

Individual-level phenotypic and genetic data for 23andMe research participants are not consented for sharing.

## ACKNOWLEDGEMENTS

We would like to thank the research participants and employees of 23andMe for making this work possible.

The following members of the 23andMe Research Team contributed to this study: Stella Aslibekyan, Adam Auton, Elizabeth Babalola, Robert K. Bell, Jessica Bielenberg, Jonathan Bowes, Katarzyna Bryc, Ninad S. Chaudhary, Daniella Coker, Sayantan Das, Emily DelloRusso, Sarah L. Elson, Nicholas Eriksson, Teresa Filshtein, Pierre Fontanillas, Will Freyman, Zach Fuller, Chris German, Julie M. Granka, Karl Heilbron, Alejandro Hernandez, Barry Hicks, David A. Hinds, Ethan M. Jewett, Yunxuan Jiang, Katelyn Kukar, Alan Kwong, Yanyu Liang, Keng-Han Lin, Bianca A. Llamas, Matthew H. McIntyre, Steven J. Micheletti, Meghan E. Moreno, Priyanka Nandakumar, Dominique T. Nguyen, Jared O’Connell, Aaron A. Petrakovitz, G. David Poznik, Alexandra Reynoso, Shubham Saini, Morgan Schumacher, Leah Selcer, Anjali J. Shastri, Janie F. Shelton, Jingchunzi Shi, Suyash Shringarpure, Qiaojuan Jane Su, Susana A. Tat, Vinh Tran, Joyce Y. Tung, Xin Wang, Wei Wang, Catherine H. Weldon, Peter Wilton, Corinna D. Wong. A.R., J.S., S.A., L.K., and K.H. are employed by and hold stock or stock options in 23andMe, Inc.

